# Exploring disease-drug pairs in Clinical Trials information for personalized drug repurposing

**DOI:** 10.1101/2023.05.04.23289463

**Authors:** Andrea Álvarez-Pérez, Lucía Prieto-Santamaría, Esther Ugarte-Carro, Belén Otero-Carrasco, Adrián Ayuso-Muñoz, Alejandro Rodríguez-González

**Affiliations:** Centro de Tecnología Biomédica, Universidad Politécnica de Madrid, Pozuelo de Alarcón, 28223, Madrid, Spain; ETS Ingenieros Informáticos, Universidad Politécnica de Madrid, Boadilla del Monte, 28660, Madrid, Spain

**Keywords:** Drug Repurposing, Clinical Trials, Precision Medicine, DISNET knowledge

## Abstract

Drug repurposing, the process of finding new uses for existing drugs, has gained considerable attention due to its potential to reduce the time and costs associated with drug development. Personalized drug repurposing, in which drugs are selected based on the characteristics of individual patients, is an emerging approach that holds promise for improving clinical outcomes. In this context, exploring disease-drug pairs in already conducted clinical trials can provide valuable insights to identify promising patient populations for further study that may lead to personalized drug repositioning. Our analysis aims to shed a light into clinical outcomes by selecting the most appropriate repurposed drug based on clinical trials patient groups’ characteristics, such as age and gender. It also gives information about the state of the clinical trials studying these disease-drug pairs, gathering information about the study type, phase and statistical method used to calculate the p-value of the chosen outcome measurement, among others. Overall, this study highlights the importance of using existing knowledge as an initial framework to facilitate further research, particularly in providing patient-specific information. Furthermore, it underlines the importance of building on previous research to facilitate a comprehensive understanding of the research topic, which can eventually improve patient outcomes.

## I. Introduction

Drug repurposing (DR) is the process of finding new therapeutic indications for existing drugs [1]. In drug discovery, DR has become a promising strategy, attracting attention to both the pharmaceutical and the research community, due to the notorious advantages over de novo drug discovery [2]. The rationale for this approach includes the reduction of the pharmaceutical research and costs, the development timelines and risk, and the use of existing data on safety and toxicity.

As a consequence of this targeted therapy, the traditional first-line drug for a disease could change, and the second- or third-line drugs could now become the first-line therapy based on the specific genetics of a patient [3]. The concept of personalized medicine is based on the premise that individuals exhibit distinct and intricate features at the molecular, physiological, environmental, and behavioural levels, necessitating customized interventions for their specific diseases that are tailored to these features [4]. Therefore, it can be stated that the aim of personalized medicine is to diagnose patients and prescribe drugs tailored to the individual’s molecular biology of their disease [1]. A clinical trial is a research study in which human volunteers are assigned to interventions based on a protocol and are evaluated for effects on biomedical or health outcomes. Currently, clinical trials study the efficacy of certain drugs to treat diseases in a certain part of the population [5].

Clinicaltrials.gov ^1^ is a web-based resource that provides easy access to information on publicly and privately supported clinical studies on a wide range of diseases and conditions [6]. It was launched as a result of the Food and Drug Administration Modernization Act of 1997 (FDAMA) and contains more than 440,000 research studies in 221 countries. With the aim of extending the usability of Clinicaltrials.gov data for aggregate analysis and public use of data for research purposes [7], the Database for Aggregate Analysis of ClinicalTrials.gov (AACT)^2^ was created, constituting an enhancement of the original database. This database is a PostgreSQL relational database storing information about clinical studies that have been registered at ClinicalTrials.gov, which content is downloaded daily.

Currently, medicine is focused on chronically treating disease symptoms, without attending to the patient profile. However, due to disease heterogeneity, this has proven to be insufficient, and causes inefficient or imprecise drug therapy and clinical trials with a high failure rate [8], [9]. Due to disease heterogeneity, prescribing drugs only to a responsive subgroup of patients would improve the cost-effectiveness of the treatment. If we are able to subclassify patients according to their molecular characteristics, candidate drugs would be more likely to succeed in clinical trials instead of appearing ineffective [1]. The number of patients who would otherwise be prescribed an ineffective drug and experience adverse effects would decrease, would then have an opportunity to undertake other therapeutic regimens that might be beneficial for them. Another fact to consider is the patient heterogeneity. Although almost the entire human genome is the same for the entire species, the presence of polymorphisms generates genotypic alterations reflected in a modification of the individual’s phenotype. If these polymorphisms are present in drug-metabolizing enzymes, drug transporters, receptors and other drug targets, this can alter the efficacy and toxicity of the drugs administered [10]. For example, ten percent of Caucasians have intermediate activity and 0.33% have no activity in this enzyme, resulting in enhanced adverse effects when taking thiopurine drugs [11].

With regard to DR, DISNET ^3^ project incorporates biomedical knowledge from structured and unstructured public sources to include data about diseases, symptoms, genes, drugs, and the relationships between them [12]. By this biomedical information integration, the project aims to provide researchers with accessible data and methodologies to better understand diseases and generate new repurposing hypotheses [2]. A set of these validated DR cases are evaluated in this study. By identifying the potential use of a drug in a new therapeutic area, and then analysing which population groups have successfully completed clinical trials for that disease-drug pair, can help lay the groundwork for more accurate analyses. By using patient-specific data and a greater number of features, personalised medicine and DR can work together to improve patient outcomes.

The main objective of this paper is to explore the current landscape of clinical trials involving drugs and diseases known to have participated in drug repurposing successful scenarios. The paper is organized as follows: Section II describes the materials and methods, including the data employed and the analyses performed. Section III details the results obtained and discusses them. And, finally, Section IV includes the conclusions and Section V identifies the limitations of the present work and the future research lines to be addressed.

## II. MATERIALS AND METHODS

We here present the process followed in the extraction of information, the processing of the data collected, and the analysis of the features of interest for the descriptive study.

### A. Information retrieval and filtering

#### 1) Disease-drug pairs

In our study, the initial dataset comprises the identification codes of the disease-drug pairs that form the basis of repurposing cases. These cases were collected from the scientific literature and mapped to DISNET’s vocabularies in Prieto Santamaría et. al [2]. Diseases are identified by the Unified Medical Language System (UMLS) [13] Concept Unique Identifier (CUI), and drugs by the CHEMBL [14] ID. Disease and drug names were extracted from DISNET database using the list of CUIs and CHEMBL IDs as input. The initial number of disease-drug pairs was 81.

We used the disease-drug names pairs to extract through Clinicaltrials.gov (CT) API all the National Clinical Trial number (NCT ID), the Condition Name, which is the primary disease being studied in the clinical trial, and the Intervention Name, which refers to the clinical intervention performed to each patient group in the study, associated to them. The API was used to download clinical trial records in a tab-separated text format as defined by the API XML schema [11]. The search query parameters used are the name of the disease and the drug in pairs. However, the ‘search expression’ parameter used in the CT wrapper searches the words at any part of the clinical trial report, including the exclusion criteria. To solve this, we employed the ‘Condition Name’ and ‘Intervention Name’ columns to cross-reference against our list of repurposed diseases and drugs, ensuring that studies outside the desired parameters did not go unnoticed. We also retained only those disease-drug pairs with at least 15 associated studies, i.e., those pairs to the right of the red line in Figure 1. For the sake of clarity, only a range between 0 to 70 number of studies is represented, although 8 more disease drug pairs are included with a number of studies superior to 70, including ‘Acquired Immunodeficiency Syndrome-Zidovudine’ and ‘HIV Infections-Zidovudine’ with 360 each. After this filtering process, from the 5,732 total retrieved studies, we kept 2,937 corresponding to 55 disease-drug pairs.

**Fig 1:**
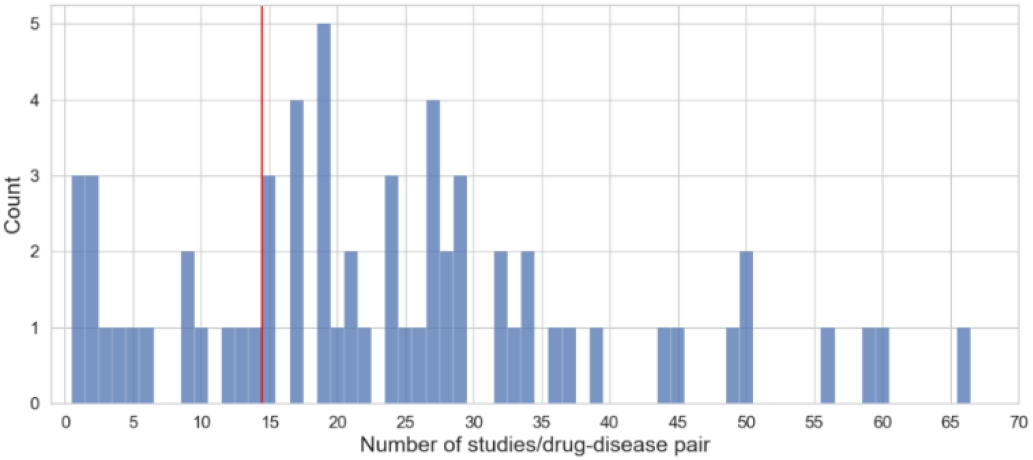
Distribution of number of disease-drug pairs associated with a discrete number of studies in a range from 0 to 70 is represented. The red line represents the threshold.

With the NCT IDs of the clinical trials, we connected to AACT relational database, which content is downloaded from clinicaltrials.gov daily. The information retrieval from AACT was done filtering by Intervention type = ‘Drug’, and the number of studies dropped to 2,876. Data from three different tables in the database were extracted:

- Studies’ table: the main table in the database contains basic information of each study, including the title, the start and completion dates, the phase of the study, and the recruitment status, among others.
- Statistics’ table: results of scientifically appropriate statistical analyses performed on primary and secondary study outcomes. It includes results for treatment effect estimates, confidence intervals and p-values.
- Eligibilities’ table: contains information about the criteria used to select participants, including inclusion and exclusion criteria.

All the steps performed in the filtering and harmonization of the data are shown in Figure 2. Some of the NCT IDs duplicated since they are associated with more than one disease-drug pair. This happened when the disease belonged to a major group of diseases, so the search expression parameter found both disease names mentioned in the study. Therefore, duplicate NCT IDs were removed regardless of its classification in the three files.

**Fig 2:**
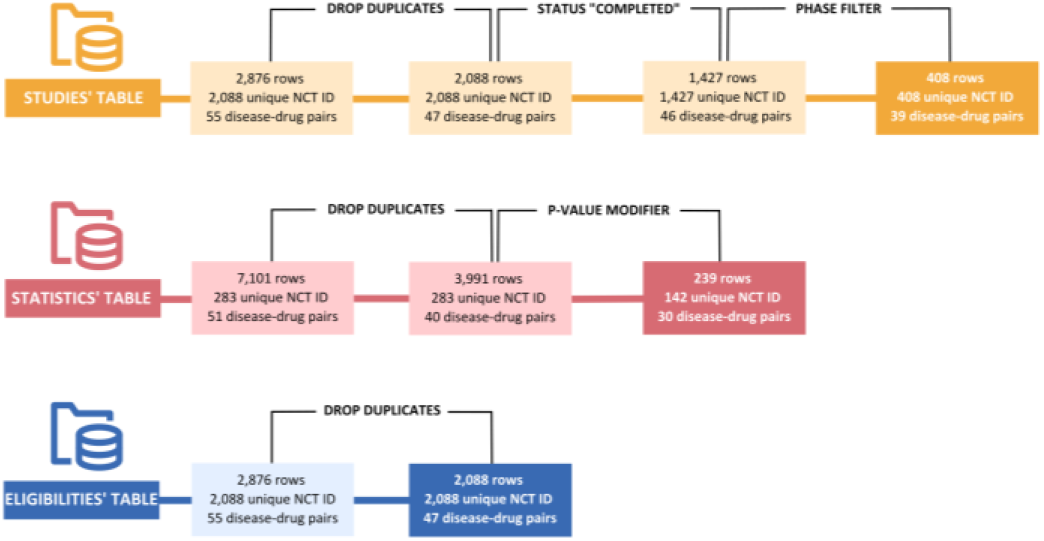
Filtering methodology flowchart.

From the studies’ table, we took only those trials that included: a current, overall status of ‘Completed’, which means the study concluded normally and participants are no longer receiving an intervention or being examined. This parameter means that the phase in which the study is located is completed. If the clinical trial is not divided into phases, the conclusion of the study is indicated by a ‘complete’ status. We also include clinical trials with an annotated phase of ‘Phase 4’ or ‘Not Applicable’. It is an exclusive feature for ‘Interventional’ type studies, where participants are assigned prospectively to an intervention according to a protocol. Typically, participation in a clinical trial requires involvement in a single phase of the study. When an interventional study is ‘Completed’ in ‘Phase 2’, it means that the activity of the participants has concluded for that specific stage of the study. Therefore, if we are interested in those studies that are close to being fully approved, we must apply these two filters jointly. We ended up with 408 unique NCT IDs corresponding to 39 disease-drug pairs. It is worth highlighting that one of the dropped disease-drug pair is ‘HIV Infections-Zidovudine’, which in previous steps was the third disease-drug pair with most studies associated. This happened due to the repeated NCT IDs with ‘Acquired Immunodeficiency Syndrome-Zidovudine’ pair. This also happens to ‘Malignant neoplasm of breast’ and ‘Breast Carcinoma’ or ‘Osteosarcoma’, and ‘Osteosarcoma of bone’.

For the statistics’ table, it should be noted that there can be more than one line per study, as multiple statistical tests can be performed. We also took only those studies in which the p-value modifier was ‘<‘. This is the standard method for conducting a hypothesis test. From this table, we only kept the NCT IDs which appeared on the studies table by an intersection of both tables. These are the clinical trials that met the conditions for status and phase. In this case, one NCT ID can be associated with more than one disease-drug pair. From the 408 unique NCT IDs of the studies table, 239 have at least one associated statistical test, and correspond to 30 disease-drug pairs.

Lastly, for the eligibilities’ table we only dropped duplicated studies. In this case, each line corresponds to a unique study with its eligibility criteria. After intersecting the file with the studies table, we only kept those NCT IDs present in the studies table. There are 39 disease-drug pairs associated to these 408 NCT IDs.

#### 2) Drugs

Initially, we collected the list of unique drug names which participate in any of the DR cases. In the 55 disease-drug pairs, there are only 29 unique drugs, corresponding to a total of 15,805 studies. The following action involved utilizing the Python wrapper of clinicaltrials.gov API to collect the NCT ID and condition name for all the studies. After removing duplicated NCT IDs, we had 14,896 studies left, which information was extracted from the AACT relational database to collect data of the studies’ table. Then, we filtered the remaining NCT IDs by the previous criteria, study status and phase, leading to a total of 2,082 studies.

#### 3) Diseases

Within the cohort of 55 disease-drug pairs analysed, a total of 47 unique diseases were identified. However, the disease entity of “malignant neoplasms” was excluded due to its broad classification and disproportionately high number of associated studies (between six- and ten-fold greater than the remaining diseases). A total of 229,221 studies were queried by the disease name using the clinicaltrials.gov API to obtain the NCT ID for each study. After removing duplicate IDs, a final dataset of 130,835 studies was retained for subsequent analysis. We extracted the AACT data for these NCT IDs and filter them by status and phase. The remaining ascend to 13,422 study count.

### B. Descriptive analyses

A comparative analysis was conducted to examine the various characteristics that define each disease-drug pair. They included study type, phase, if it uses a FDA regulated drug, the statistical method used to perform evaluate the p-value of the analysis, the age and gender of the participants, and if they are affected or not by the disease. In most cases, the comparison was carried out through graphical representation of the study count according to the different variables analysed in order to identify patterns. Tables were also used to compare other variables in a more detailed way.

Furthermore, we also compared the distributions of the study count between disease-drug pairs and the entirety of the information hosted at the AACT database in order to determine whether DR cases followed different distributions from the general trends.

## III. RESULTS AND DISCUSSION

All the files and procedures carried out are available at: https://medal.ctb.upm.es/internal/gitlab/disnet/personalized-dr

### 1) Study type

According to clinicaltrials.gov, the study type refers to the nature of the investigational use for which the clinical study information is being submitted. In ‘Interventional’ studies, participants are assigned to an intervention according to a protocol to evaluate the effect of the intervention on biomedical or health-related outcomes. This kind of study type is dominant in most of the studies referring to the disease-drug pairs studied for repositioning, and exclusive for 16 disease-drug pairs. On the contrary, in ‘Observational’ studies, the outcomes are assessed in pre-defined groups of individuals and the investigator does not assign specific interventions to the study participants. This kind of study is exclusive for 4 of the disease-drug pairs. The rest of them count with both types of studies.

### 2) Study phase

This attribute is specific only to those studies of the ‘Interventional’ type. A study which is completed in ‘Phase 4’ means that the study was divided into phases and has passed all of them. ‘Not Applicable’ is used to describe trials without FDA-defined phases. This type of structuring is common in studies of devices or behavioral interventions, but it does not rule out that the treatment of said study is pharmacological therapy. ‘Observational’ studies that lack this feature have been classified separately as ‘Observational’. Behavioral interventions are designed to affect the actions that individuals take regarding their health. This can be applied to study as the ultimate goal of the study, the psychological response of the patient and different therapies.

An example of a particular study would be ‘*NCT01161017: Mechanism-based Choice of Therapy for Pain: Can Successful Prevention of Migraine be Coupled to a Psychophysical Pain Modulation Profile?*’. In this study, two different pharmacological therapies are used, Amitriptyline and Topiramate, to compare the response at the psychophysical level of patients with migraine. Figure 3 shows the study count per disease-drug pair classified by the three possible structure a study can present, mentioned above. It can be said that:

**Fig 3:**
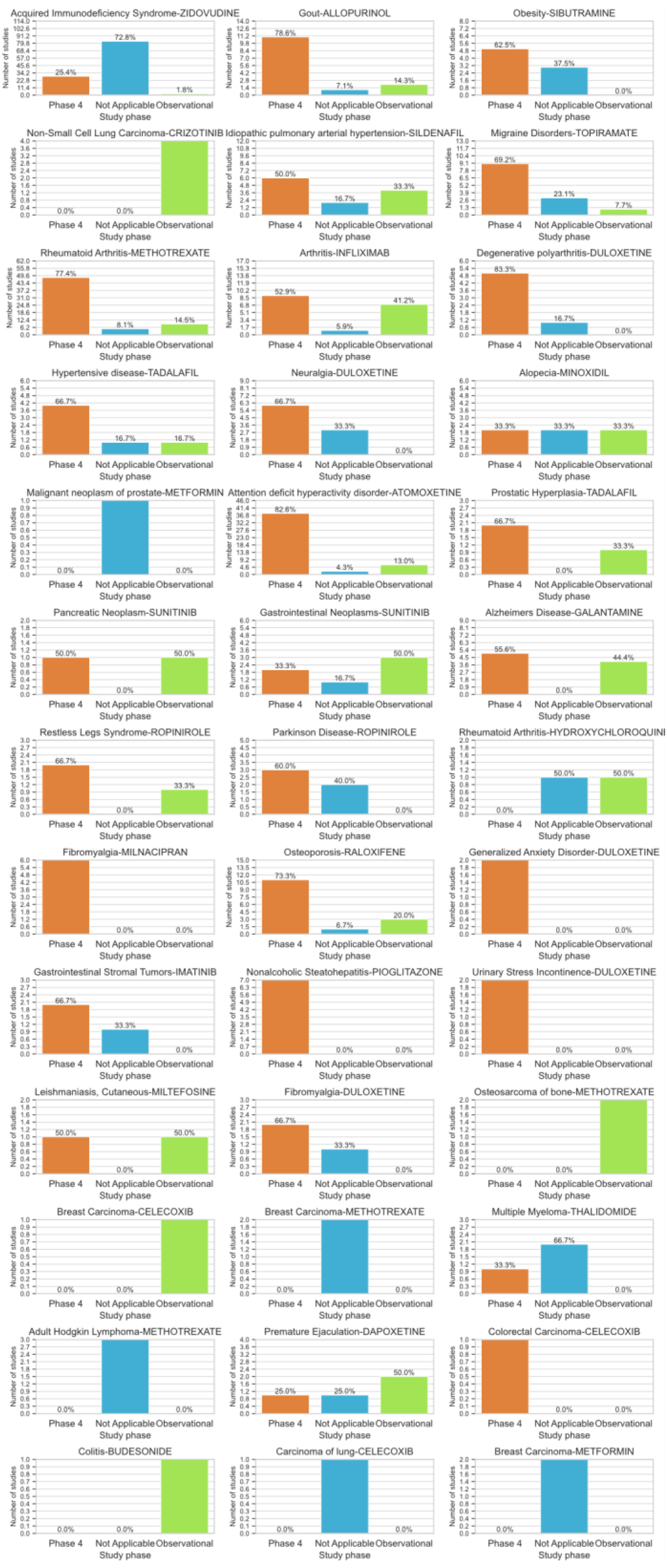
Phases distribution per disease-drug pair. Interventional type studies are the sum of ‘Phase 4’ and ‘Not Applicable’ classes.

- 5 out of the 16 ‘Interventional’-exclusive disease-drug pairs are only divided into phases.
- 5 out of the 16 ‘Interventional’-exclusive disease-drug pairs have only ‘Not Applicable’ studies, although the number of studies per pair is between 1 and 3.
- The trend is to combine studies of both types, and in these cases the percentage of ‘Phase 4’ type studies is higher in the great majority of cases.
- The percentage of studies labeled as ‘Observational’ corresponds to the assignment of ‘Observational’ type studies in the graph representing the ‘Study type’. At the end, this chart presents a breakdown of the ‘Interventional’ study type column from the previous section.

### 3) Accepts healthy volunteers

In general, 90.44% of the studies in our subset do not accept healthy volunteers. There is not any disease-drug pair whose studies use healthy or sick volunteers exclusively in the 100% of the cases. Disease-drug pairs with a considerable number of studies that accept healthy volunteers, taking as thresholds (1) a number greater than 3 and (2) a percentage of the total study greater than 20%, would narrow to ‘Obesity - Sibutramine’ pair.

### 4) Age intervals

Although almost all disease-drug pairs have been studied for similar age intervals in patients, we can highlight ‘Attention deficit hyperactivity disorder - Atomoxetine’, which is usually studied in patients inside narrower age ranges, mainly separating young groups from adults and elder patients. This also happens with ‘Idiopathic pulmonary arterial hypertension - Sildenafil’, which is normally divided into patients taking approximately 20 years as a cutoff. There are also larger studies for this disease-drug pair, where the minimum age ranges from zero months or days, up to an indefinite maximum age in years.

### 5) Statistical method

All p-values associated to all statistical tests performed were < 0.05. The proportion of each of the statistical methods used by disease-drug pair is depicted in Figure 4. The mean of statistical tests’ methods used per disease-drug pair is two, and it depends on the number of studies performed. The distribution of the methods is heterogeneous, a pattern or trend is not appreciated. However, it can be stated that an increase in the number of studies performed leads to an increase in the number of methods used to calculate the p-value of the clinical trial. This means there is no any guideline stablished in terms of calculating the statistical significance in clinical trials yet.

**Fig 4:**
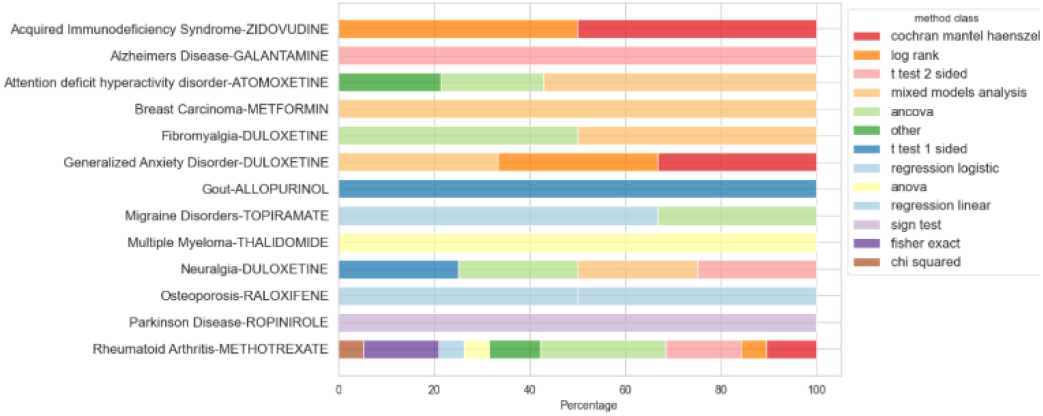
Percentages of the statistical methods used per disease-drug pair.

### 6) Gender-based studies

Most of the studies accept all gender participants. Table I only shows those disease-drug pairs with at least one gender-based study. ‘Prostatic Hyperplasia’, ‘Premature ejaculation’ or ‘Malignant neoplasm of prostate’ refer to male class, and ‘Breast Carcinoma’, ‘Urinary stress incontinence’ and ‘Osteoporosis’ to female participants. ‘Breast Carcinoma - Methotrexate’ pair presents a study which gender is defined as ‘All’, but the study is: ‘A Pilot Study in Women with Operable Breast Cancer’.

**TABLE I.** GENDER-BASED STUDIES DISTRIBUTION.

| Disease-drug | Gender | 'Gender-based' count | 'All gender' study count |
| --- | --- | --- | --- |
| Gout-Allopurinol | Male | 2 | 12 |
| Obesity-Sibutramine | Female | 1 | 7 |
| Acquired Immunodeficiency Syndrome-Zidovudine | Male/Female | 7 | 107 |
| Rheumatoid Arthritis-Methotrexate | Female | 1 | 61 |
| Neuralgia-Duloxetine | Female | 1 | 8 |
| Alopecia-Minoxidil | Male/Female | 5 | 1 |
| Prostatic Hyperplasia-Tadalafil | Male | 3 | - |
| Malignant neoplasm of prostate-Metformin | Male | 1 | - |
| Attention deficit hyperactivity disorder-Atomoxetine | Female | 1 | 45 |
| Gastrointestinal Neoplasms-Sunitinib | Female | 1 | 5 |
| Osteoporosis-Raloxifene | Male/Female | 14 | - |
| Fibromyalgia-Milnacipran | Female | 2 | 4 |
| Gastrointestinal Stromal Tumors-Imatinib | Male | 1 | 2 |
| Urinary Stress Incontinence-Duloxetine | Female | 2 | - |
| Fibromyalgia-Duloxetine | Female | 1 | 2 |
| Breast Carcinoma-Methotrexate | Female | 1 | 1 |
| Breast Carcinoma-Celecoxib | Female | 1 | - |
| Premature Ejaculation-Dapoxetine | Male | 4 | - |
| Breast Carcinoma-Metformin | Female | 2 | - |

### 7) Comparative analysis of diseases and drugs

After intersecting both DR and CT filtered drug datasets, we kept 26 unique drug names. In Figure 5, we analyzed the distribution of the number of studies corresponding to each drug. Green bars correspond to the left y-axis, which measures the study count for all CT database. Pink bars measure the study count for our DR drug cases and are related to the right y-axis. The graph does not show a clear relationship between the number of total studies associated with each drug and the number of studies using that drug as a repositioning case. We have to take into account that the y-axis scale in the ‘CT dataset’ is 5 times the ‘DR dataset’ scale. In general, the number of studies corresponding to the ‘DR dataset’ accounts for a small proportion of the ‘CT dataset’, especially for Metformin. Only for Atomoxetine, Zidovudine and Methotrexate is the proportion slightly higher. These drugs are the most studied as replacement cases in proportion to the number of total studies.

**Fig 5:**
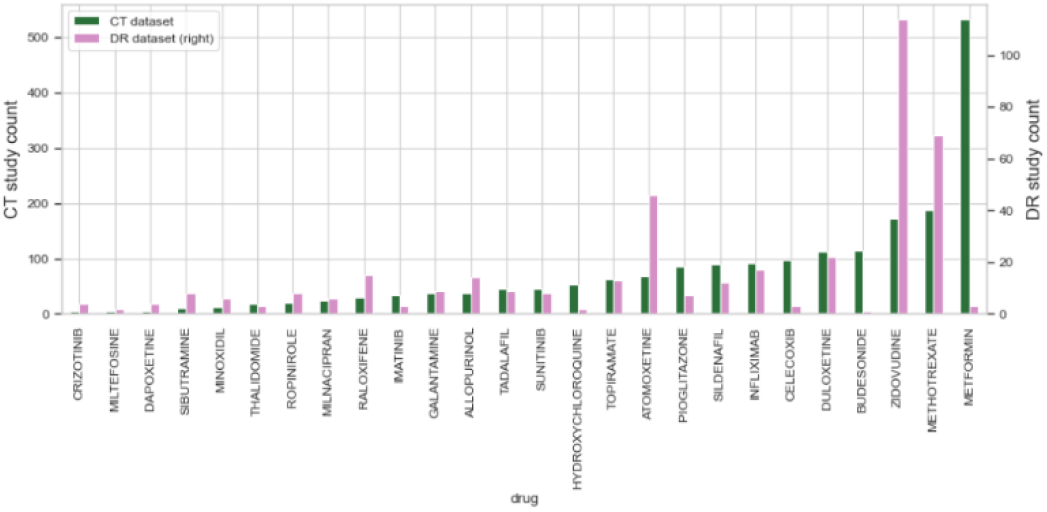
Study count per drug in both ‘CT dataset’ (green) and ‘DR dataset’ (pink)

In the case of the diseases, we also intersected the DR and CT datasets, keeping 32 unique disease names. Figure 6 represents the distribution of the number of studies corresponding to each dataset and disease. Red bars correspond to the whole CT database (left y-axis) and light blue bars for the DR diseases study count (right y-axis). This case is slightly similar as the drugs. The y-axis scale in the ‘CT dataset’ is this time 40 times the ‘DR dataset’, further accentuating the difference in the number of studies devoted to each disease overall versus the number of studies devoted to each disease when it is part of a drug repositioning case. The only case with a slightly higher proportion is ‘Attention deficit disorder’, without being appreciable. This may happen because studied diseases are very general and encompass a large number of subtypes that can be treated with different drugs.

**Fig 6:**
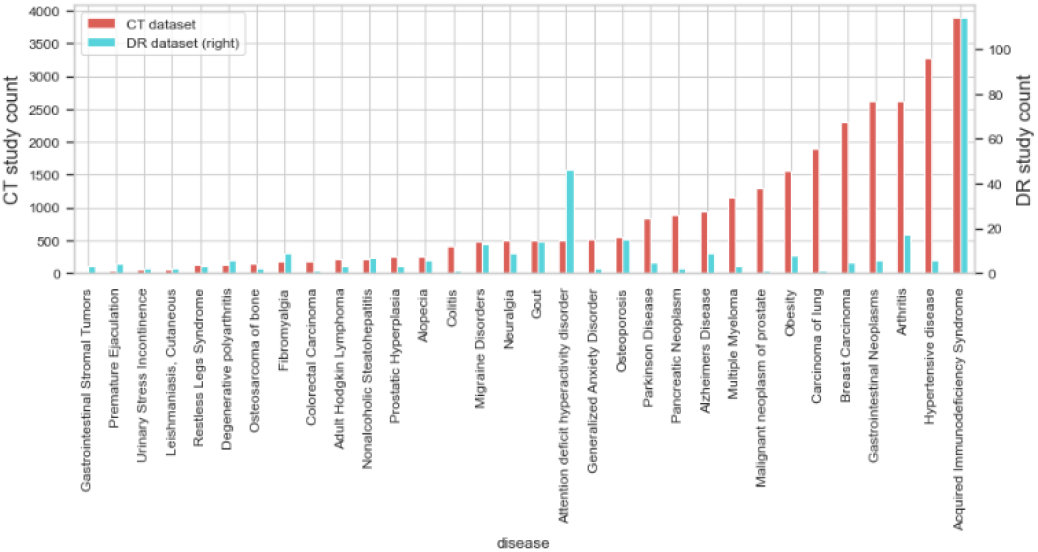
Study count per disease in both ‘CT dataset’ (red) and ‘DR dataset’ (blue)

## IV. CONCLUSIONS

Human diseases are heterogeneous and complex. Similar to how personalized medicine is useful for treating individual patients based on their unique characteristics, personalized DR offers new opportunities to tailor particular patients’ needs. Clinicaltrials.gov and the AACT provide an extensive database of publicly and privately supported clinical studies, which can be used for aggregate analysis and research purposes. The present study involves conducting a comprehensive analysis of successfully completed studies for each disease-drug pair. The objective is to identify the pairs that have yielded a higher success rate in studies conducted on patient cohorts categorized by age and gender. Additionally, this study seeks to determine which types of studies and statistical methods are more likely to yield positive outcomes. Furthermore, we compared the total number of registered studies (successfully completed) in AACT with the number of studies dedicated to DR cases defined for each disease and drug individually. This allows us to determine the approximate proportion of total studies dedicated to investigating the feasibility of drug repositioning.

## V. LIMITATIONS AND FUTURE WORK

The limitations of this study include the lack of information more specific patient information. We have information about groups of patients, but not for each of them individually. Further research is needed to explore the potential of drug repurposing in personalized medicine and to identify new repurposable drugs for the treatment of various diseases. There are several potential future directions for this paper. First, the current analysis could be expanded to include more databases and drug-repositioned pairs. Furthermore, incorporating additional features such as drug-drug interactions, adverse drug reactions, and pharmacokinetic properties could provide a more comprehensive understanding of the potential use of repurposed drugs. Finally, it would also be interesting to investigate the regulatory barriers that could hinder the approval of repurposed drugs, as well as the economic and social impact of such drug.

## Data Availability

All data produced are available online at

https://medal.ctb.upm.es/internal/gitlab/disnet/personalized-dr

## Acknowledgment

This research was funded by the project “Data-driven drug repositioning applying graph neural networks (3DR-GNN)” (PID2021-122659OB-I00) from the Spanish Ministerio de Ciencia e Innovación and “Drug repurposing hypotheses through a data-driven approach (GRENADA)” (PDC2022-133173-I00) from the Spanish Ministerio de Ciencia e Innovación and MadridDataSpace4Pandemics, funded by Comunidad de Madrid (Consejería de Educación, Universidades, Ciencia y Portavocía) with FEDER funds as part of the response from the European Union to COVID-19 pandemia. Andrea Álvarez Pérez was granted by Universidad Politécnica de Madrid and Banco Santander for a predoctoral ‘Programa Propio’ grant. Belen Otero Carrasco’s work is supported by “Formación de Personal Investigador” grant (FPI PRE2019-090912) as part of the project “DISNET (Creation and analysis of disease networks for drug repurposing from heterogeneous data sources applied to rare diseases)” (RTI2018-094576-A-I00) from the Spanish Ministerio de Ciencia e Innovación.

## Footnotes

1 https://clinicaltrials.gov/

2 https://ctti-clinicaltrials.org/

3 https://disnet.ctb.upm.es/

## Notes

### Competing Interest Statement

The authors have declared no competing interest.

### Funding Statement

This study was funded by the project 'Data-driven drug repositioning applying graph neural networks (3DR-GNN)' (PID2021-122659OB-I00) from the Spanish Ministerio de Ciencia e Innovación and 'Drug repurposing hypotheses through a data-driven approach (GRENADA)' (PDC2022-133173-I00) from the Spanish Ministerio de Ciencia e Innovación and MadridDataSpace4Pandemics, funded by Comunidad de Madrid (Consejería de Educación, Universidades, Ciencia y Portavocía) with FEDER funds as part of the response from the European Union to COVID-19 pandemia. Andrea Álvarez Pérez was granted by Universidad Politécnica de Madrid and Banco Santander for a predoctoral 'Programa Propio' grant. Belén Otero Carrasco's work is supported by 'Formación de Personal Investigador' grant (FPI PRE2019-090912) as part of the project 'DISNET (Creation and analysis of disease networks for drug repurposing from heterogeneous data sources applied to rare diseases)' (RTI2018-094576-A-I00) from the Spanish Ministerio de Ciencia e Innovación.

